# Impact of an Integrated Intervention Package During Preconception, Pregnancy, and Early Childhood on inflammation, IGF-1, IGFBP3 during first 6 months of life: Findings from the WINGS Randomized Controlled Trial

**DOI:** 10.1101/2025.08.10.25333400

**Authors:** Ranadip Chowdhury, Urmimala Maiti, Sarita Devi, Roshni M. Pasanna, Sunita Taneja, Partha P Majumder, Tor A. Strand, R M Pandey, Anura V Kurpad, Souvik Mukherjee, Nita Bhandari

**Affiliations:** Society for Applied Studies, 45, Kalu Sarai, New Delhi, India; DBT and Wellcome India Alliance Clinical and Public health Fellow, Hyderabad, India; Department of Physiology, St. John’s Medical College, Bengaluru, India; Human Genetics Unit, Indian Statistical Institute, Kolkata, West Bengal, India; Department of Research, Innlandet Hospital Trust, Lillehammer, Norway; Department of Biostatistics, All India Institute of Medical Sciences, New Delhi, India; Human Microbiome Research Laboratory, Biotechnology Research Innovation Council-National Institute of Biomedical Genomics, Kalyani, West Bengal, India; Regional Centre for Biotechnology, NCR Biotech Science Cluster, Faridabad, India

## Abstract

**Background:** Early-life interventions targeting maternal and child health, nutrition, and psychosocial care may influence infant growth and immune development by modulating systemic inflammation and growth factor pathways. However, causal evidence of comprehensive, integrated interventions initiated during early life is limited.

**Methods:** The study was nested in a factorial randomized controlled trial (RCT) in women aged 18–30 years. Participants included in this study were randomized to receive either a preconception intervention package or routine care until early childhood. The intervention included health care for growth-related conditions, nutrition, WASH (water, sanitation, and hygiene), and psychosocial care. The primary study demonstrated a substantial effect of the intervention on growth and development. Blood samples from 381 (178 from the intervention group and 203 from the control group) infants were analyzed at 3 and 6 months of age for inflammatory and growth-related biomarkers; C-reactive protein (CRP), alpha-1-acid glycoprotein (AGP), insulin-like growth factor 1 (IGF-1), and IGF binding protein 3 (IGFBP3). Generalized linear models were used to estimate the mean differences and relative risks of inflammatory markers and growth-related biomarkers between the intervention and routine care groups. This trial was registered at Clinical Trials Registry–India: CTRI/2020/10/028770.

**Results:** Baseline maternal characteristics were similar between the two study groups. At 3 and 6 months, the proportion of infants with CRP levels greater than 5 mg/L were similar in both groups. No significant differences were observed in AGP or IGF-1 concentrations at either time point. IGFBP3 was lower in the intervention group at 3 months (adjusted mean difference: –30.8 ng/mL; 95% CI: –55.3, –6.3), but this effect was not sustained at 6 months.

**Conclusions:** An integrated intervention delivered from preconception through pregnancy and early childhood did not result in reductions in markers of systemic inflammation or sustained improvements in growth factor profiles during the first six months of life. These findings highlight the complexity of early-life inflammatory processes and underscore the need for further research on the long-term and synergistic effects of early interventions in low-resource settings.

## Introduction

The continuum from preconception through pregnancy and early childhood constitutes a critical window for influencing growth, development, and long-term health outcomes. Substantial evidence suggests that environmental exposures and targeted health and nutrition interventions during these critical periods can have lasting effects on metabolic programming, immune system maturation, and developmental trajectories. (1,2,3) Of particular interest are the complex interplay between systemic inflammation (indicated by raised C-reactive protein or alpha-1-acid glycoprotein levels) and key growth mediators such as insulin-like growth factor 1 (IGF-1) and its principal binding protein, IGFBP3, given their integrated roles in somatic growth, neurodevelopment, immune regulation, and susceptibility to disease.(4,5) Persistent low-grade inflammation in early life has been linked to suboptimal growth and an increased risk of non-communicable diseases (NCDs). In contrast, a diverse and resilient gut microbiome, together with physiologically appropriate concentrations of IGF-1 and IGFBP3, is essential for supporting normal linear growth and optimal neurodevelopment. (6) Interventions addressing maternal and child nutrition, infection control, and psychosocial stimulation have demonstrated the potential to modulate these biological pathways and improve child health outcomes positively. (7) There is a paucity of research evaluating the effects of comprehensive, integrated intervention strategies initiated preconceptionally and sustained through pregnancy and early childhood on inflammatory profiles and growth factor dynamics in infants. To address this gap, we recently conducted the Women and Infants Integrated Growth Study (WINGS), an individually randomized factorial trial designed to assess the impact of a multidomain intervention package including health, nutrition, psychosocial care and support, and water, sanitation, and hygiene (WASH) delivered from preconception through early childhood. (8) The primary outcomes demonstrated that this integrated intervention significantly reduced the incidence of low birth weight (LBW), small for gestational age (SGA), and stunting at 24 months compared to standard care.

Building on these findings, the present study aimed to investigate the effects of the WINGS intervention on inflammatory markers and IGF-1/IGF-BP3 concentrations during the first six months of life. By addressing maternal and infant health, nutrition, and environmental exposures during critical developmental windows, the intervention may foster a favorable gut microbial milieu, attenuate systemic inflammation, and optimize growth factor profiles in infancy. (9) We hypothesized that the integrated approach implemented in the WINGS trial would result in reduced inflammation and improved levels of IGF-1 and IGF-BP3 in the intervention group compared to the group receiving routine care. This study aims to enhance our understanding of the mechanisms by which early-life interventions in low-resource settings modulate inflammation and growth factor axes, providing valuable insights into the biological pathways that link these interventions to improved growth and developmental outcomes.

## Materials and methods

### Primary Study design

This was a factorial randomized controlled trial (RCT) conducted in low- and middle-income neighbourhoods of Delhi, India. Eligible women of reproductive age (18-30 years), identified through door-to-door survey were enrolled and randomized to receive either the preconception intervention package or routine care. They were followed for 18 months. If they were identified as pregnant in this period (ultrasonographic confirmation of pregnancy), they were then randomized either to receive the pregnancy and early childhood intervention package or routine care. All pregnant women were followed until delivery, and their children until they reached 24 months of age. Data of those children were included in analysis who reached 24 months of age by 30^th^ June 2021.The primary study has been detailed elsewhere by Taneja et al. (8) This two-step randomization resulted in four groups: preconception and pregnancy interventions (A), preconception interventions only (B), pregnancy and early childhood interventions only (C), and no preconception interventions, with routine pregnancy and early childhood care (D). The interventions, under the four domains of health, nutrition, WASH, and psychosocial care and support, are briefly described below.

In the pre-conception period, women were screened and treated for medical conditions, including anemia. All received one tablet of iron and folic acid (IFA) daily and weekly multiple micronutrients (MMN) supplements to meet ½ to ¾ of the recommended daily allowance (RDA). They also received an albendazole tablet twice a year. All women were counseled on adequate diets, positive thinking, problem-solving skills, and menstrual and hand hygiene.

### Intervention group

In the pregnancy and early childhood interventions, women received monthly antenatal care, screening, and treatment for medical conditions, including anemia. Women received daily IFA, MMN (∼1 RDA), calcium, and vitamin D throughout pregnancy. Albendazole was given once during pregnancy. Women with BMI<25 kg/m^2^ in the second and third trimesters received food supplements. Women with inadequate weight gain (IWG) were identified based on the Institute of Medicine guidelines for gestational weight gain (GWG), and nutritional counseling and extra food supplements were provided.(8) Water filters, water storage bottles, hand washing stations, soap, and disinfectants were provided to all families, in addition to counseling on sanitation and hygiene. Women received counselling on exclusive breastfeeding during the first 6 months of the infant’s age. Additional visits were made for babies born preterm, LBW, and for mothers with breastfeeding problems. Nutritional supplementation included vitamin D for all infants, iron for very low and low birth weight infants, and daily snacks and supplements for mothers. Complementary food supplements (milk-cereal mix) were started at six months and continued until 24 months. Mothers were taught age-specific child play, responsive care, and stimulation activities. Weights were measured during home visits, and children with inadequate weight gain were referred to lactation counselors and pediatricians.

### Control group

Women in the control group were advised to seek care from government sources (free of cost) to access family planning services and weekly iron folic acid supplementation. They were advised to register for antenatal care at a government or private facility, and have at least four antenatal care checkups, consume iron folic acid, calcium, vitamin D daily throughout pregnancy, access supplementary foods through the Integrated Child Development Services (ICDS) scheme and plan to deliver in health facilities. Women were advised to go for a postnatal health check-up and to consume iron, folic acid, calcium, vitamin D, and supplementary foods daily through the ICDS scheme. Mothers were advised to breastfeed their babies exclusively for the first six months and continue breastfeeding for at least two years. They were also encouraged to arrange home visits by the community health workers in the first 42 days of life.

### Participants of the current study

Between 1^st^ December 2020 and 30^th^ December 2021, 381 mother-infant dyads were selected consecutively when the infants completed 6 months of age from Group A and D—as defined in the primary WINGS study. Randomization was conducted by an independent statistician, who provided a list to ensure unbiased group allocation. All outcome assessments were conducted by a separate team that was not involved in the intervention delivery and was blinded to group assignments before the measurement process.

Ethical clearance for this study was granted by the Ethics Review Committee of the Society for Applied Studies in New Delhi, India. The study complied with ethical standard outlined in Declaration of Helsinki. This study was registered with the Clinical Trial Registry of India under the identifier CTRI/2020/10/028770. Written informed consent was secured primarily from mothers before their inclusion in the study.

### Sample Size

Few intervention studies have systematically examined the infant blood markers of inflammation and growth during the first six months of life. In the WINGS study, we hypothesized a 0.2 standard deviation (SD) increase in Length-for-Age Z-score (LAZ) and a 25% relative reduction in the incidence of stunting at 24 months in the group receiving interventions during preconception, pregnancy, and early childhood (Group A), compared to the group receiving routine care (Group D) (5). For the present study, we hypothesized that the intervention package would result in increased IGF-1 levels and reduced inflammatory markers, such as CRP, with an anticipated effect size equal to or greater than that observed in the WINGS study. Assuming a corresponding 0.3 SD decrease of CRP in the intervention group with the power at 80% and 95% confidence level, we calculated that a total of 352 infant blood samples would be required —176 in the intervention group and 176 in the routine care group.

### Sample collection and laboratory analysis

We collected 3 mL of blood from infants at 3 and 6 months of age. The samples were transported from the field to the “Clinical and Research laboratory” in SAS at 4°C on ice packs and centrifuged to extract serum. The serum was then stored at −80°C for analysis of IGF-1, IGBP3, CRP, and AGP.

Serum IGF-1 and IGFBP3 were analyzed by electrochemiluminescence immunoassay on the Cobas e 601 analyzer (Roche Diagnostics, Mannheim, Germany) using the Elecsys IGF-1 kit (Roche Diagnostics, Ref No. 07475896190), and IGFBP-3 kit (Roche Diagnostics, Ref No. 07574690190), according to the manufacturer’s instructions. Calibration and internal quality controls were performed using Roche-provided calibrators and control materials. Results were expressed in nanograms per milliliter (ng/mL). Intra and Inter assay CVs were <3 and <4% respectively.

Serum CRP and AGP were measured by immunoturbidometric based assay on the Cobas c 501 analyzer (Roche Diagnostics, Mannheim, Germany). All reagents, calibrators, and controls were obtained from Roche Diagnostics, and the assay was performed according to the manufacturer’s guidelines. Quality control procedures were performed using standard control materials supplied by the manufacturer. Results were expressed in milligrams per liter (mg/L) and grams per liter (g/L) for CRP and AGP, respectively. All samples were analyzed in a single batch to reduce inter-assay variability. Intra- and inter-assay CVs were <2% and <4%, respectively.

### Analysis Plan

Baseline characteristics of the preconception, pregnancy, and early childhood group (Group A) and the routine care group (Group D) were compared using means and standard deviations for continuous variables, and proportions for categorical variables. All analyses followed an intention-to-treat approach and were conducted using STATA version 16. Generalized linear models (GLMs) with a Gaussian family and identity link function were used to estimate mean differences in CRP, AGP, IGF-1, and IGFBP3 concentrations. To estimate risk ratios for inflammatory status between infants in the intervention and routine care groups, GLMs with a binomial family and log link function were employed. Final models were adjusted for place of birth.

## Results

### Baseline characteristics

Between December 2020 and December 2021, 381 infants (178 in the preconception, pregnancy, and early childhood intervention group, and 203 in the routine care group) were recruited for this study (Figure 1). The baseline characteristics of the women were similar, except for the proportion of underweight women, the number of families possessing a below the poverty line (BPL) card, and the place of birth (Table 1).

**Table 1.** Baseline characteristics of mother at recruitment.

|  | Preconception, pregnancy<br>and early childhood<br>(Group A)<br>(n=178) | Routine care<br>(Group D)<br>(n=203) |
| --- | --- | --- |
| Woman age (years) | 24.02 (2.99) | 23.99 (3.09) |
| Woman height (cm) | 151.99 (5.27) | 152.40 (5.75) |
| BMI category (kg/m<sup>2</sup>); n (%) |  |  |
| ≥25 | 44 (24.72) | 48 (23.64) |
| 18.5 - 24.99 | 107 (60.11) | 118 (58.13) |
| <18.5 | 27 (15.17) | 37 (18.23) |
| Joint/ Extended family; n (%) | 109 (61.24) | 127 (62.56) |
| Women schooling >12 yrs; n (%) | 103 (57.87) | 107 (52.71) |
| Homemaker; n (%) | 174 (97.75) | 195 (96.06) |
| Family has BPL card; n (%) | 6 (3.37) | 16 (7.88) |
| Family covered by health insurance scheme; n (%) | 11 (6.18) | 20 (9.85) |
| Place of delivery; n (%) |  |  |
| Large hospital | 124 (69.66) | 68 (33.50) |
| Middle level hospital/ small hospital/ birthing centre | 50 (28.09) | 116 (57.14) |
| Home | 4 (2.25) | 19 (9.36) |

\*2 twins 1 in Preconception, pregnancy and early childhood group and 1 in Routine care group; n (%) unless specified otherwise; BPL= Below Poverty Line; All values expressed as Mean (SD) unless specified otherwise.

**Figure 1.**
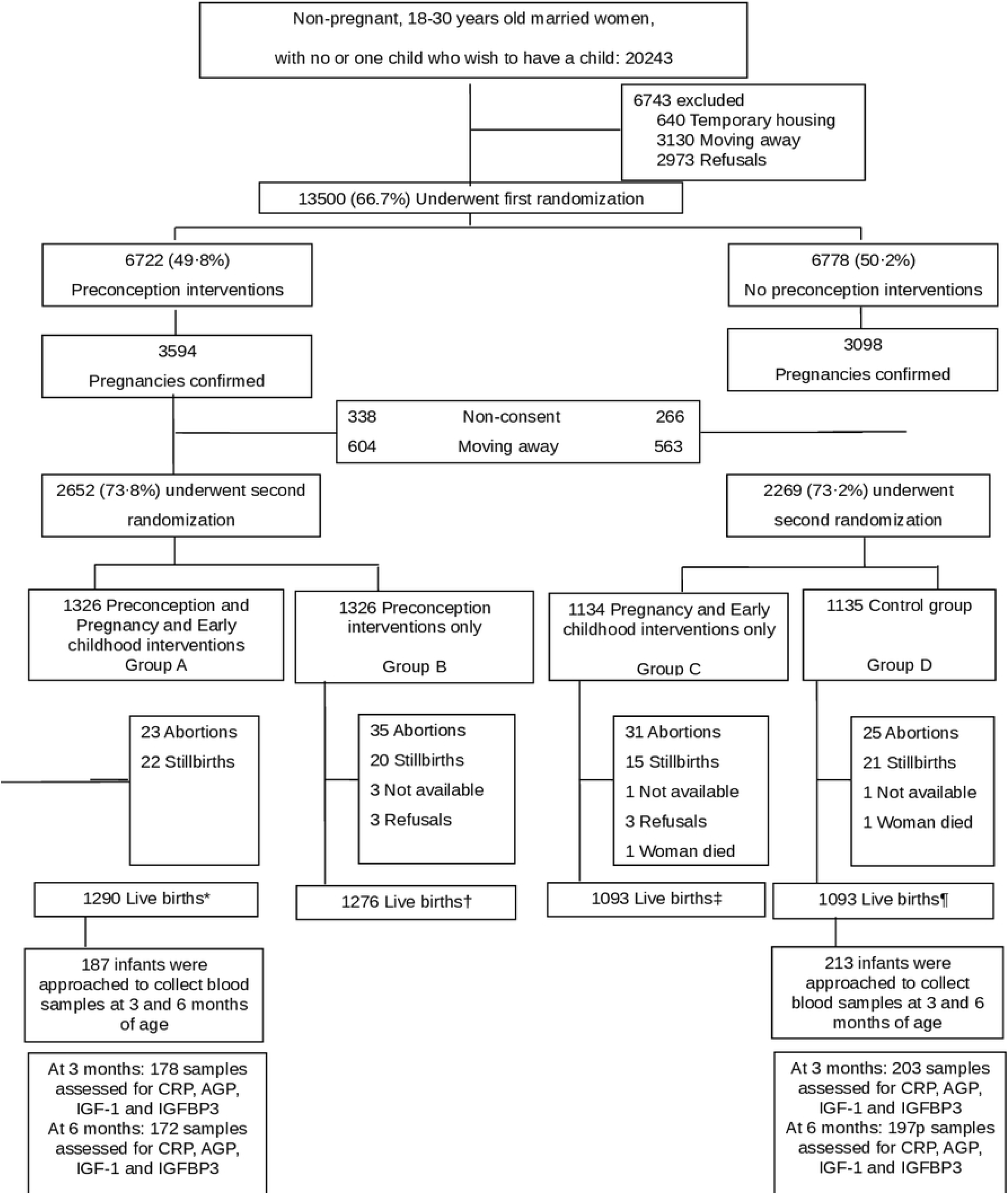
Screening, enrollment, randomization and follow up.

Table 2 compares infant inflammatory and growth-related biomarker levels during the first 6 months of life between the Preconception, Pregnancy, and Early Childhood group and the Routine Care group. CRP, AGP, and IGF-1 levels were similar between the groups at both 3 and 6 months. IGFBP3 levels at 3 months were lower in the Preconception, pregnancy, and early childhood group compared to the Routine care group (adjusted mean difference: −32.49 ng/mL; 95% CI: −56.79, −8.19), but this difference was not sustained at 6 months.

**Table 2.** Infant inflammatory growth biomarkers levels during the first 6 months of life.

| Inflammatory markers and growth-related hormones | Preconception, pregnancy | Routine care (Group D) (n=203) | Unadjusted mean difference | Adjusted mean difference |
| --- | --- | --- | --- | --- |

|  | and early<br>childhood<br>(Group A)<br>(n=178) |  |  |  |
| --- | --- | --- | --- | --- |
| CRP at 3 months (mg/L) | 2.20 (7.64) | 3.16 (9.78) | -0.96 | -0.65 |
| Mean (SD) |  |  | (-2.74, 0.82) | (-2.55, 1.25) |
| Median (IQR) | 0.59 (0.58,<br>0.71) | 0.59 (0.58,<br>1.06) |  |  |
| CRP at 6 months<sup>1</sup> (mg/L) | 2.42 (7.62) | 1.66 (4.13) | 0.76 | 1.03 |
| Mean (SD) |  |  | (-0.47, 1.99) | (-0.29, 2.36) |
| Median (IQR) | 0.59 (0.58,<br>0.66) | 0.59 (0.58,<br>0.59) |  |  |
| AGP at 3 months (g/L) |  |  |  |  |
| Mean (SD) | 0.55 (0.24) | 0.57 (0.29) | -0.02<br>(-0.07, 0.03) | -0.02<br>(-0.08, 0.03) |
| Median (IQR) | 0.5 (0.36,<br>0.64) |  | 0.5 (0.35, 0.7) |  |
| AGP at 6 months<sup>1</sup> (g/L) |  |  |  |  |
| Mean (SD) | 0.58 (0.38) | 0.56 (0.29) | 0.02<br>(-0.05, 0.08) | 0.01<br>(-0.06, 0.08) |
| Median (IQR) | 0.46 (0.37,<br>0.66) | 0.49 (0.38,<br>0.65) |  |  |
| IGF-1 at 3 months (ng/mL) |  |  |  |  |
| Mean (SD) | 9.84 (4.90) | 10.03 (5.29) | -0.19<br>(-1.22, 0.84) | -0.36<br>(-1.46, 0.75) |
| Median (IQR) | 6.99 (6.98, 11.21) | 6.99 (6.98, 11.7) |  |  |
| IGF-1 at 6 months<sup>†</sup> (ng/mL) |  |  |  |  |
| Mean (SD) | 8.94 (4.40) | 9.26 (4.48) | -0.32<br>(-1.23, 0.59) | -0.30<br>(-1.29, 0.67) |
| Median (IQR) | 6.99 (6.98, 8.69) | 6.99 (6.98, 8.83) |  |  |
| IGFBP3 at 3 months (ng/mL) |  |  |  |  |
| Mean (SD) | 181.20<br>(114.70) | 210.04 (109.95) | -28.84<br>(-51.60, -6.09) | -32.49<br>(-56.79, -8.19) |
| Median (IQR) | 172 (78, 273) | 222 (112, 303) |  |  |
| IGFBP3 at 6 months (ng/mL) |  |  |  |  |
| Mean (SD) | 1523.42<br>(497.45) | 1496.46<br>(475.81) | 26.95<br>(-72.72, 126.63) | -5.16<br>(-112.28, 101.95) |
| Median (IQR) | 1504 (1259, 1764) | 1499.5 (1201.5, 1800.5) |  |  |

Note: Values are means ± SDs (n) or mean differences (95% CI); Adjustment was done for place of delivery;

Table 3 compares the prevalence of elevated inflammatory markers in infants during the first six months of life between the Preconception, Pregnancy, and Early Childhood group and the Routine Care group. The elevated inflammatory status was similar at 3 and 6 months of life in both the Preconception, Pregnancy, and Early Childhood group and the Routine Care group. The intervention group had a lower proportion of infants with CRP >5 mg/L compared to the routine care group at 3 months. However, this effect was attenuated by adjusting for potential confounders. (adjusted RR = 0.49; 95% CI: 0.22, 1.09).

**Table 3.** Infant inflammatory status during first 6 months of life.

| Inflammatory markers | Preconception,<br>pregnancy and<br>early childhood<br>(Group A)<br>(n=178) | Routine care<br>(Group D)<br>(n=203) | Unadjusted RR<br>(95% CI) | Adjusted RR<br>(95% CI) |
| --- | --- | --- | --- | --- |
| CRP > 5 (mg/L) at 3<br>months | 9 (5.06) | 22 (10.84) | 0.47 (0.22, 0.98) | 0.49 (0.22, 1.09) |
| CRP >5 (mg/L) at 6<br>months | 19 (10.67) | 18 (8.87) | 1.20 (0.65, 2.22) | 1.18 (0.61, 2.28) |
| AGP > (g/L) at 3<br>months | 11 (6.18) | 13 (6.40) | 0.96 (0.44, 2.10) | 0.92 (0.40, 2.14) |
| AGP > (g/L) at 6<br>months | 21 (11.80) | 20 (9.85) | 1.20 (0.67, 2.22) | 1.1 (0.60, 2.10) |

Adjusted for place of delivery,

## Discussion

In this randomized controlled trial, we evaluated the impact of an integrated intervention package delivered from preconception through pregnancy and early childhood on infant inflammatory markers and growth-related hormones during the first six months of life. We found that CRP, AGP, and IGF-1 levels were similar between the intervention and routine care groups at both 3 and 6 months. IGFBP3 levels at 3 months were lower in the intervention group compared to the routine care group, but this difference was not sustained at 6 months. These results suggest that, despite the comprehensive nature of the intervention, which included nutritional supplementation, infection control, WASH improvements, and psychosocial support, there was no consistent or sustained effect on systemic inflammation or growth factor profiles in early infancy. (11,12) Several factors may explain the lack of differences observed in our study. First, the prevalence of raised inflammatory markers in this population was relatively low, which may have limited the potential for further reduction through intervention. Second, the timing and duration of the intervention, while spanning critical developmental windows, may not have been sufficient to elicit measurable changes in systemic biomarkers within the first six months of life. It is also possible that the benefits of the intervention on inflammation and growth factors may emerge later in childhood, as has been observed in some longitudinal studies. (14) Third, the multifactorial etiology of early-life inflammation, including genetic, environmental, and infectious contributors, may attenuate the impact of even comprehensive intervention packages. The transient reduction in IGFBP3 at 3 months in the intervention group warrants further exploration. Previous studies have demonstrated a negative correlation between inflammatory markers and growth-related hormones, i.e., a rise in CRP and AGP corresponded with a decrease in IGF-1 and IGFBP3 levels. (4) IGFBP3 is the primary binding protein for IGF-1, playing a crucial role in regulating IGF-1 bioavailability and activity. While lower IGFBP3 could theoretically enhance IGF-1 action, we did not observe corresponding increases in IGF-1 concentrations at this early time point. This case control study in Zimbabwe found that IGF-1 and IGFBP-3 decreased with time from 3 months of age among both stunted and non-stunted children. (4) The biological significance of this isolated finding in our study is, therefore, unclear and may reflect normal and substantial physiological variation. A prospective cohort study on healthy infants examining the effect of nutritional differences on growth factor levels found that dairy-based protein intake was associated with increased levels of both IGF-1 and IGFBP-3 between 9 and 12 months of age, compared to non-dairy animal source protein intake. (15) Given the complexity of our intervention, the observed effects are likely due to the combined influence of multiple components, making it difficult to attribute changes to any single factor. Additionally, conclusions based solely on data from 3 and 6 months may be inappropriate.

Further research is needed to translate the data on IGF-1 and IGFBP-3 into meaningful diagnostic outcomes, considering the influence of age, gender, and genetic factors, among other determinants.

Our study has several strengths, including its randomized design, rigorous implementation of a multidomain intervention, and careful adjustment for potential confounders such as place of delivery, socioeconomic status, and maternal BMI. The use of standardized laboratory methods and blinded outcome assessment further enhances the validity of our findings. However, some limitations should be acknowledged. The sample size, while adequate for detecting moderate effect sizes, may have been insufficient to identify smaller but clinically meaningful differences. Additionally, the study was conducted in a single urban setting in India, which may limit generalizability to other populations. (13)

## Conclusion

Our findings suggest that an integrated intervention package delivered from preconception through early childhood do not reduce systemic inflammation or improve growth factor profiles during the first six months of life. These results highlight the complexity of early-life inflammatory processes and suggest that additional or alternative strategies may be necessary to achieve meaningful improvements in these biomarkers, as well as the consideration of other biomarkers that reflect inflammatory responses. Our study contributes to the growing body of evidence on the challenges and opportunities of early-life interventions in low-resource settings.

## Data Availability

Data Availability Statement: Please find the dataset DOI: 10.5061/dryad.6t1g1jxbs

http://datadryad.org/share/8L0Hg_iTK6InXiR2pME1kTVDY3Yv0dQjdBVAU2U4s8c

## Acknowledgments

We deeply acknowledge the contribution of the participants and their families and are thankful to the community leaders for their cooperation.

## Data sharing

Individual requests will also be considered on a case-by-case basis. The request for data should be accompanied by a detailed proposal describing the scientific questions to be addressed.

Proposals should be submitted to RC.

## Author contribution

RC was responsible for study design, implementation and development of the data management system, data analysis; UM was responsible for data analysis; SD and RM were responsible for all laboratory analysis and quality control; RC, UM drafted the manuscript and prepared tables and figures. PPM, AVK, TS, RMP, ST, SM, NB provided technical guidance and all authors: read and approved the final manuscript.

## Clinical Trial Registry number and website

This trial was registered at Clinical Trials Registry–India as CTRI/2020/10/028770; https://ctri.nic.in/Clinicaltrials/advsearch2.php

## References

1. Colombo J, Gustafson KM, Carlson SE. Critical and sensitive periods in development and nutrition. Annals of Nutrition and Metabolism. 2019 Dec 28;75(Suppl. 1):34–42.

2. Koletzko B, Brands B, Demmelmair H, Early Nutrition Programming Project. The Early Nutrition Programming Project (EARNEST): 5 y of successful multidisciplinary collaborative research. The American journal of clinical nutrition. 2011 Dec 1;94:S1749–53.

3. Vaivada T, Gaffey MF, Bhutta ZA. Promoting early child development with interventions in health and nutrition: a systematic review. Pediatrics. 2017 Aug 1;140(2).

4. Prendergast AJ, Rukobo S, Chasekwa B, Mutasa K, Ntozini R, Mbuya MN, Jones A, Moulton LH, Stoltzfus RJ, Humphrey JH. Stunting is characterized by chronic inflammation in Zimbabwean infants. PloS one. 2014 Feb 18;9(2):e86928.

5. Suchdev PS, Namaste SM, Aaron GJ, Raiten DJ, Brown KH, Flores-Ayala R, BRINDA Working Group. Overview of the biomarkers reflecting inflammation and nutritional determinants of anemia (BRINDA) project. Advances in Nutrition. 2016 Mar 1;7(2):349–56.

6. Ronan V, Yeasin R, Claud EC. Childhood development and the microbiome—the intestinal microbiota in maintenance of health and development of disease during childhood development. Gastroenterology. 2021 Jan 1;160(2):495–506.

7. Tessema TT, Alamdo AG, Mekonnen EB, Yirtaw TG, Debele FA, Gemechu T, Belachew T. The effect of psychosocial stimulation on the development, nutrition, and treatment outcomes of hospitalised children with severe acute malnutrition in Southern Ethiopia: a cluster randomised control trial: EPSoSAMC study. Journal of Nutritional Science. 2025 Jan;14:e17

8. Taneja S, Chowdhury R, Dhabhai N, Upadhyay RP, Mazumder S, Sharma S, Bhatia K, Chellani H, Dewan R, Mittal P, Bhan MK. Impact of a package of health, nutrition, psychosocial support, and WaSH interventions delivered during preconception, pregnancy, and early childhood periods on birth outcomes and on linear growth at 24 months of age: factorial, individually randomised controlled trial. Bmj. 2022 Oct 26;379.

9. Nigam M, Devi K, Coutinho HD, Mishra AP. Exploration of gut microbiome and inflammation: A review on key signalling pathways. Cellular Signalling. 2024 Mar 16:111140.

10. National Research Council Committee to Reexamine IOMPWG. The National Academies Collection: Reports funded by National Institutes of Health. In: Rasmussen KM, Yaktine AL, eds. Weight Gain During Pregnancy: Reexamining the Guidelines. Washington (DC). National Academies Press, 2009.

11. Bhutta ZA, Bang P, Karlsson E, Hagenäs L, Nizami SQ, Söder O. Insulin-like growth factor I response during nutritional rehabilitation of persistent diarrhoea. Archives of Disease in Childhood. 1999 May 1;80(5):438–42.

12. Yang I, Corwin EJ, Brennan PA, Jordan S, Murphy JR, Dunlop A. The infant microbiome: implications for infant health and neurocognitive development. Nursing research. 2016 Jan 1;65(1):76–88.

13. Tarrade A, Panchenko P, Junien C, Gabory A. Placental contribution to nutritional programming of health and diseases: epigenetics and sexual dimorphism. Journal of Experimental Biology. 2015 Jan 1;218(1):50–8.

14. DeBoer MD, Elwood SE, Platts-Mills JA, McDermid JM, Scharf RJ, McQuade ET, Jatosh S, Houpt ER, Mduma E. Association of Circulating Biomarkers with Growth and Cognitive Development in Rural Tanzania: A Secondary Analysis of the Early Life Interventions in Childhood Growth and Development In Tanzania (ELICIT) Study. The Journal of Nutrition. 2023 May 1;153(5):1453–60.

15. Kittisakmontri K, Lanigan J, Wells JC, Manowong S, Kaewarree S, Fewtrell M. Quantity and source of protein during complementary feeding and infant growth: evidence from a population facing double burden of malnutrition. Nutrients. 2022 Sep 23;14(19):3948.

